# A healthful plant-based diet is associated with higher physical and mental well-being among older adults independent of circulating CRP

**DOI:** 10.1101/2023.11.30.23299231

**Authors:** Kerstin Schorr, Marian Beekman, Venetka Agayn, Jeanne H.M. de Vries, Lisette C.P.G.M. De Groot, P. Eline Slagboom

**Affiliations:** Section of Molecular Epidemiology, department of Biomedical Data Sciences, Leiden University Medical Center, Leiden, The Netherlands; Innoso BV, The Hague, Netherlands; Division of Human Nutrition and Health, Wageningen University, Wageningen, The Netherlands

**Keywords:** plant-based diet, subjective well-being, older adults, inflammation, dietary pattern

## Abstract

Plant-based diets (PBD) have been found to be environmentally sustainable as well as beneficial for health. Observational research showed that higher plant-based diet quality improves well-being in adult women, however this is unclear for older adults. This association may be due to anti-inflammatory properties of PBD. Older adults, often suffering from chronic inflammation, may therefore profit from a more PBD. Therefore, we investigated the relation between PBD and well-being in older adults of both genders and tested whether the effects are influenced by circulating high-senstivity C-reactive protein (hsCRP) levels. We used the data of the population-based Lifelines Cohort Study (n=6,635, mean age=65.2 years) and a subsample in which hsCRP was measured (n=2,251, mean age=65.2 years). We applied a plant-based diet index measuring adherence to a healthful (hPDI) and an unhealthful (uPDI) plant-based diet based on food frequency questionnaires. The RAND-36 questionnaire was applied as measure of quality of life, from which we derived physical (PCS) and mental component scores (MCS). Older adults with the highest adherence to a healthful plant-based diet had respectively 14% and 12% greater odds for high physical well-being and mental well-being. Meanwhile, higher adherence to uPDI was associated with respectively 19% and 14% lower odds for high physical and mental well-being. We observed an additive but no mediating effect of hsCRP on the association between plant-based diets and well-being. We conclude that in older men and women, adherence to a healthful plant-based diet and circulating levels of inflammation are independently associated with physical and mental well-being.

## Introduction

Plant-based diets are becoming increasingly popular, and dietary organizations recommend a shift towards a more plant-based diet [1-3]. The growing interest may partly be due to the fact that a plant-based diet contributes less to greenhouse gas emissions and thus to climate change [3]. Presumed benefits of healthful plant-based diets include a reduced risk for age-related diseases, such as metabolic diseases and cognitive impairment but also well-being [4-7]. With advancing age chronic illnesses and multi-morbidity become more prevalent. This highlights the benefits of adopting a plant-based diet for quality of life and mental and physical well-being beyond the effects on health.

Well-being has been associated with various different lifestyle or sociodemographic factors in older adults. Female sex, higher age, physical inactivity and the prevalence of chronic conditions have been associated with poor well-being [8]. In contrast, a healthful plant-based diet showed a positive effect on middle-aged healthy women [7]. There could however also be potential drawbacks in following plant-based diets. A meta-analysis suggested a link between adherence to a vegetarian diet and an increased risk for depression, especially among younger adults [9]. In addition plant-based diets may be accompanied by inadequate protein intake, due to lower nutritional quality of plant proteins, increasing the risk for sarcopenia and potentially contributing to lower well-being among older adults [10, 11].

An added benefit of plant-based diets may lie in their anti-inflammatory properties [12-14]. Chronic inflammation is prevalent especially among older adults and may negatively affect their well-being [15, 16]. It has been hypothesized that the association between a healthful plant-based diet and higher well-being may be due to an anti-inflammatory effect of a plant-based diet [7]. Despite this hypothesis, a mediating effect of circulating inflammatory markers on well-being in older adults has not been investigated yet. Current literature, however, has focused mainly on the effects of plant-based diets in younger adults, usually devoid of chronic inflammation, and mainly in women [17].

Therefore, it needs to be elucidated whether chronic inflammation and adherence to healthful plant-based diets have an independent effect on mental and physical well-being both in older men and women.

Here we investigate the relation between adherence to a plant-based diet and wellbeing in men and women above 60 years and whether such associations are independent of chronic inflammation established by circulating hsCRP. To answer these questions we used data from the population-base cohort Lifelines [18]. Adherence to a plant-based diet is quantified by applying a plant-based diet index (PDI) differentiating between a healthful (hPDI) and unhealthful (uPDI) plant-based diet [19], well-being is quantified by the RAND-36 questionnaire from which we derived a physical (PCS) and a mental component score (MCS), and circulating levels of high-sensitivity C-reactive protein (hsCRP) are taken as a proxy of inflammation level. Consequently, we will assess 1) how adherence to a healthful and unhealthful plant-based diet is associated with physical and mental well-being in older adults above 60 years of age and 2) whether this relationship is mediated by circulating CRP levels as proxy of inflammation levels.

## Material and Methods

### Study population

The current study investigated data from the Lifelines Cohort Study [18]. The Lifelines Cohort study is a multi-disciplinary prospective population-based cohort study examining the health and health-related behaviors of 167,729 persons living in the North of the Netherlands in a unique three-generation design. It employs a broad range of investigative procedures in assessing the biomedical, socio-demographic, behavioral, physical and psychological factors which contribute to the health and disease of the general population, with a special focus on multi-morbidity and complex genetics. The LifeLines Cohort Study is conducted according to the principles of the Declaration of Helsinki and in accordance with research code UMCG and is approved by the medical ethical committee of the University Medical Center Groningen, The Netherlands [18]. Baseline assessments were carried out from 2007 to 2013, followed by a second (2014-2017) and third (2019-2023) assessment round, interspersed with follow-up questionnaires.

For the purpose of this study we included participants, from whom food frequency questionnaire (FFQ) data, as well as quality of life data (assessed by RAND-36 questionnaire) was available. We primarily focused on older adults above 60 years of age (n=6,635, Table 1) and investigated whether similar associations could be observed in adults aged 18-60 years (N=35,850, Suppl. Table 2). To assess the relationship with circulating high sensitivity C-reactive protein levels, a subgroup of participants with available hsCRP levels were included (n_older adults_=2,251, n_younger dults_=14,699).

**Table 1.** Baseline Characteristics of Lifelines participants (>60 years) per tertiles of healthful and unhealthful PDI.

|  | Unhealthful plant-based diet index |  |  | Healthful plant-based diet index |  |  |
| --- | --- | --- | --- | --- | --- | --- |
| **N<sub>total</sub>** | **6635** |  |  | **6635** |  |  |
|  | **low** | **medium** | **high** | **low** | **medium** | **high** |
| **hPDI [18-90]** | 60.0±5.9 | 56.0±6.0 | 52.4±6.2 | 49.1±3.5 | 56.5±1.7 | 64.0±3.6 |
| **uPDI [18-90]** | 44.3±3.2 | 50.9±1.4 | 57.8±3.4 | 54.2±5.7 | 50.0±5.4 | 47.1±5.5 |
| **n** | 2598 | 2016 | 2021 | 2233 | 2218 | 2184 |
| **n<sub>females</sub> (%)** | 1444 (55.6) | 1022 (50.7) | 996 (50.7) | 1095 (49.0) | 1166 (52.6) | 1201 (55.0) |
| **Age, mean±SD** | 65.0 ± 4.6 | 65.4±4.9 | 65.2±4.8 | 65.3±4.8 | 65.1±4.7 | 65.2±4.9 |
| **BMI [kg/m<sup>2</sup>], mean±SD** | 26.6±3.7 | 26.5±3.5 | 26.4±3.5 | 26.9±3.6 | 26.5±3.4 | 26.0±3.6 |
| **Physical Activity (h/week), mean±SD** | 7.0±7.5 | 6.7±7.5 | 6.5±7.4 | 6.3±7.2 | 6.9±7.7 | 7.1±7.5 |
| **Medication use yes, n (%)** | 1701 (65.5) | 1317 (65.3) | 1317 (65.2) | 1508 (67.5) | 1409 (63.5) | 1418 (64.9) |
| **Monthly income, n (%)** |  |  |  |  |  |  |
| (**<1500€**) | 346 (13.3) | 313 (15.5) | 404 (20.0) | 383 (17.2) | 335 (15.1) | 345 (15.8) |
| (**1500-3000€**) | 1459 (56.2) | 1173 (58.2) | 1160 (57.4) | 1267 (56.7) | 1282 (57.8) | 1243 (56.9) |
| (**>3000€**) | 793 (30.5) | 530 (26.3) | 457 (22.6) | 583 (24.1) | 601 (27.1) | 596 (27.3) |
| **Physical CS, mean±SD** | 52.1±7.0 | 51.6±7.5 | 51.4±7.5 | 51.5±7.6 | 51.9±7.0 | 51.9±7.3 |
| **Mental CS, mean±SD** | 53.7±6.9 | 53.5±7.2 | 53.0±7.4 | 53.2±7.5 | 53.5±6.9 | 53.5±7.0 |
| **Including hsCRP (n=2251)** |  |  |  |  |  |  |
| **hsCRP [mg/L], mean±SD** | 2.4±4.7 | 2.4±4.5 | 2.6±4.5 | 2.7±5.1 | 2.2±3.5 | 2.3±4.7 |

hPDI=healthful plant-based diet index, uPDI=unhealthful plant-based diet index, BMI=body mass index, Physical CS=physical component score, Mental CS=mental component score, hsCRP=high-sensitivity C-reactive protein

**Table 2.** Associations between a healthful (hPDI) and unhealthful (uPDI) plant-based diet index with physical (PCS) and mental (MCS) component scores among older adults (>=60 years)

|  | Physical component score |  | Mental component score |  |
| --- | --- | --- | --- | --- |
| **hPDI** | OR [95% CI] | p | OR [95% CI] | p |
| **Tertile 1** | ref |  | ref |  |
| **Tertile 2** | 1.05 [0.93; 1.19] | 0.403 | 1.03 [0.92; 1.16] | 0.609 |
| **Tertile 3** | 1.12 [0.99; 1.27] | 0.082 | 1.14 [1.01; 1.28] | 0.043 |
| **uPDI** |  |  |  |  |
| **Tertile 1** | ref |  | ref |  |
| **Tertile 2** | 0.94 [0.84; 1.07] | 0.351 | 0.94 [0.83; 1.06] | 0.298 |
| **Tertile 3** | 0.81 [0.72; 0.92] | <0.001 | 0.87 [0.78; 0.98] | 0.027 |

n=6635, adjusted for age, gender, BMI, physical activity, medication, income and energy intake (OR 1.00, p≤0.001 in all models)

### Plant-based diet index

We applied a plant-based diet index to quantify the adherence to a plant-based diet and quality of foods consumed. The plant-based diet index (PDI) was adapted from previous publications and calculated from FFQ data [19]. The FFQ in the Lifelines Cohort Study was implemented in the form of a flower FFQ, meaning it was split into smaller questionnaires and spread between assessments [20-22]. FFQ items were classified into food groups, as displayed (Suppl. Table 1). Categorization of food items was based on previous publications and recommendations of the Netherlands nutrition center (Voedingcentrum^1^). In contrast to the original index [19], intake of certain margarines was included in the vegetable oil category, in accordance with current recommendations on healthy fats.

The intake in each food group (in g/d) was then divided into cohort-specific quintiles and points awarded per quintile. For the healthful PDI (hPDI) scale healthful plant based food groups were scored positively (1=lowest quintile, 5=highest quintile), while less healthy food groups were reverse-scaled (1=highest quintile, 5=lowest quintile). This procedure was reversed for the unhealthful PDI (uPDI), so that healthy plant foods were scored reversely, while less healthy plant foods were scored positively. Animal food groups were always scored reversely, with the lowest quintile receiving 5 points and the highest quintile 1 point. This results in indices ranging from 18-90, with higher index scores indicate higher adherence to the respective dietary pattern. As such, the score allows for comparison of plant-based diets of varying composition. To account for differences in energy intake, the indices were adjusted for overall energy-intake using the residual method, by regressing energy intake on intake per food group [23].

### Well-being

Well-being was measured by the RAND-36 questionnaire, a tool to assess quality of life in eight subdomains, covering areas of both physical and mental health [24]. Data was available in a subgroup of people that participated in the Lifelines baseline assessment (2007-2013). Subdomain scores were derived according to scoring instructions^2^. From the scores of the subdomains, physical and mental components scores were calculated by standardizing the scores using population-specific factors for the Dutch population. In a second step, to calculate the aggregated physical and mental component scores, the standardized subdomain scales are multiplied by their respective factor coefficient and summed. In a last step, the component scores are normalized, resulting in a score in which a mean of 50 (and standard deviation of 10) represents the mean of the Dutch population. The computation of the component scores and population-specific factors are described in detail elsewhere [25, 26]. For the purpose of this study, the component scores were binarized by the median, in order to allow for comparison of subjects with low and high well-being.

### Inflammatory marker

Blood samples were collected via venopuncture before 10 am in the morning in fasted participants between 2008 and 2013. High-sensitivity CRP was measured in heparin using kits from two different suppliers, either by CardioPhase hsCRP, Siemens Healthcare Diagnostics, Marburg, Germany or CRPL3, Roche Diagnostics, Mannheim, Germany. Measurements were carried out at the Groningen University Medical Center [18].

### Covariates

In the statistical analyses, we made adjustments for covariates. Age, gender, body mass index (BMI), physical activity, income, use of prescription medication and energy intake were considered as covariates. We included prescription medication as it is a good indicator for disease prevalence, which is likely related to both CRP and well-being. Income has also been found to be associated with well-being in previous research [27]. Information on age, gender, BMI, physical activity, income and use of prescription medication was derived from questionnaires [18]. Physical activity was measured by the Squash questionnaire [28] and reported in minutes/week of moderate and vigorous physical activity [29]. To obtain a more interpretable unit we converted this physical activity variable into hours/week. Use of prescription medication and income was self-reported by participants. Net income was categorized as low (<1500€), medium (1500€ to 3000€) and high (>3000€/month). Since a slight increase in total energy intake across tertiles of hPDI was observed, energy intake was additionally adjusted for.

### Statistical Analysis

To assess the association between the physical and mental component scores and the plant-based diet indices, a logistic regression with binarized component scores as the outcome was applied. The PDIs were treated as predictor variables and divided into tertiles, while adjusting for age, gender, BMI, physical activity, income, use of prescription medication and energy intake. In the first step, models were calculated separately for older (>60 years) and adults aged 18-60 and stratified by gender (Model 1, Suppl. Material 1). Secondly, to assess a potential mediation by circulating hsCRP, the association between hsCRP and plant-based diet adherence was assessed via linear regression, with hsCRP as outcome and PDIs as predictor variables, while adjusting for covariates. In the last step of the mediation analysis, hsCRP was added to the models of the first step (Model 2, Suppl. Material 1). As sensitivity analysis, the association between an overall plant-based diet index and well-being was assessed using the logistic regression model from the first step (Model 3, Suppl. Material 1). Further, we replicated our findings by applying a linear regression with rank-inverse normal transformed well-being scores as the outcome. All analyses were carried out in R.

## Results

### Sample Characteristics

Among older adults, above 60 years, those with a high adherence to a healthful plant-based diet (hPDI), i.e. those in the highest tertile of hPDI were more likely to be female, had a lower BMI and were slightly more physically active than those in the lowest tertile of hPDI. Further, those with the highest adherence to hPDI were less likely to take prescription medication and to have a low income. Meanwhile those with a high adherence to an unhealthful plant-based diet, i.e. in the highest tertile of uPDI were more likely male, less physically active and had a lower income as compared to those in the lowest tertile of uPDI (Table 1). Similar differences across tertiles were observed among younger adults (Suppl. Table 2). Among younger adults, those in the highest tertile of hPDI had higher intakes of protein, particularly plant protein, fiber and unsaturated fatty acids compared to those in the highest tertile of uPDI (Suppl Table 3).

### Plant-based diet index associated with well-being

When comparing extreme tertiles among older adults, (n=6,635) above 60 years of age, those with high adherence to a healthful plant-based diet had 14% greater odds for high mental well-being (OR 1.14, p=0.043) and 12% greater odds for high physical well-being (OR 1.12, p=0.082). Those with the highest adherence to an unhealthful plant-based diet had 19% lower odds for high physical well-being (OR 0.81, p=<0.001) and 13% lower odds for high mental well-being (OR 0.87, p=0.027), after adjusting for covariates. (Table 2). In younger adults (n=35,850), between 18 and 60 years, similar associations of adherence to the PDIs and with physical and mental well-being were observed (Table 3). High adherence to a healthful plant-based diet was significantly associated with 17% higher odds of high physical well-being (OR=1.17, p<0.001) and 8% greater odds for high mental well-being (OR 1.08, p=0.01). A high adherence to an unhealthful plant-based diet was associated with 16% lower odds for high physical well-being (OR 0.84, p<0.001) and 14% lower odds for high mental well-being (OR=0.86, p=<0.001).

**Table 3.** Associations between an unhealthful plant-based diet index (uPDI) and physical (PCS) and mental (MCS) component scores among young adults (<60 years)

|  | Physical component score |  | Mental component score |  |
| --- | --- | --- | --- | --- |
| **hPDI** | OR [95% CI] | p | OR [95% CI] | p |
| **Tertile 1** | ref |  | ref |  |
| **Tertile 2** | 1.10 [1.05; 1.16] | <0.001 | 1.03 [0.98; 1.08] | 0.240 |
| **Tertile 3** | 1.17 [1.10; 1.24] | <0.001 | 1.08 [1.02; 1.14] | 0.010 |
| **uPDI** |  |  |  |  |
| **Tertile 1** | ref |  | ref |  |
| **Tertile 2** | 0.92 [0.88; 0.97] | 0.002 | 0.91 [0.86; 0.96] | <0.001 |
| **Tertile 3** | 0.84 [0.79; 0.89] | <0.001 | 0.86 [0.82; 0.91] | <0.001 |

n=35850, adjusted for age, gender, BMI, physical activity, medication, income and energy intake

### Associations of hPDI and uPDI with mental and physical component scores similar in men and women

To explore whether the associations between PDIs and physical and mental component sores in equally present in men and women, we performed stratified analysis. The association of hPDI with both PCS and MCS seem only marginally stronger in women. The association of uPDI with mental well-being seemed to be stronger in women (OR 0.78, p=0.008) than in men (OR 0.92, p=0.314), while the association of uPDI with PCS is similar between older men and women (Table 4). Further, among younger adults, the association of hPDI with both, PCS and MCS, appeared to be stronger among men. Mostly no differences were found between men and women in the association between uPDI adherence and physical and mental component (Suppl. Table 4). However, because of the overlapping confidence intervals, there is no clear evidence for differences in association between men and women.

**Table 4.** Associations between an unhealthful plant-based diet index (uPDI) with physical (PCS) and mental (MCS) component scores stratified by gender among older adults (>60 years)

|  | Physical component score |  | Mental component score |  |
| --- | --- | --- | --- | --- |
|  | OR [95% CI] | p | OR [95% CI] | p |
| **Females** |  |  |  |  |
| **hPDI Tertile 1** |  |  |  |  |
| **hPDI Tertile 2** | 1.08 [0.92; 1.28] | 0.354 | 1.07 [0.91; 1.25] | 0.424 |
| **hPDI Tertile 3** | 1.16 [0.97; 1.38] | 0.105 | 1.16 [0.98; 1.38] | 0.083 |
| **Males** |  |  |  |  |
| **hPDI Tertile 1** |  |  |  |  |
| **hPDI Tertile 2** | 1.10 [0.93; 1.31] | 0.274 | 1.02 [0.86; 1.20] | 0.863 |
| **hPDI Tertile 3** | 1.11 [0.92; 1.33] | 0.269 | 1.11 [0.93; 1.32] | 0.245 |
| **Females** |  |  |  |  |
| **uPDI Tertile 1** | ref |  | ref |  |
| **uPDI Tertile 2** | 0.93 [0.79; 1.10] | 0.404 | 0.91 [0.77; 1.06] | 0.231 |
| **uPDI Tertile 3** | 0.87 [0.73; 1.04] | 0.134 | 0.78 [0.67; 0.94] | 0.008 |
| **Males** |  |  |  |  |
| **uPDI Tertile 1** | ref |  | ref |  |
| **uPDI Tertile 2** | 0.98 [0.82; 1.17] | 0.835 | 0.97 [0.82; 1.15] | 0.736 |
| **uPDI Tertile 3** | 0.84 [0.71; 1.01] | 0.058 | 0.92 [0.77; 1.09] | 0.314 |

Adjusted for age, gender, BMI, physical activity, medication, income and energy intake

### Plant-based diest and circulating CRP levels have additive effects on mental and physical component score

Previous research indicated an association of CRP with plant-based diet intake and with decreased well-being [14, 16]. Therefore, we aimed to assess whether circulating CRP levels mediate the association of plant-based diets and well-being. Mean hsCRP was slightly higher in older adults compared to younger adults (Table 1, Suppl. Table 2). We first assessed whether hsCRP is associated with uPDI and hPDI in the Lifelines study population. In older adults, we observed that higher adherence to hPDI was significantly associated with lower circulating hsCRP (p=0.0025 for medium and p=0.057 for high adherence). Meanwhile, higher adherence to uPDI was associated with higher levels of circulating hsCRP, in younger but not older adults (p<0.001, Suppl. Table 5).

HsCRP was available in n=2251 older adults together with dietary and well-being data. In this smaller dataset high adherence to hPDI was not significantly associated with physical (OR 1.13, p=0.258) and mental well-being (OR=1.06, p=0.548) anymore, with a considerable drop in effect noticeable for mental well-being. Similarly, uPDI was not significantly associated with PCS (OR 0.84, p=0.10) and with MCS (OR 0.87, p=0.171) anymore, likely due to limited power. Given the fact that direction of effects was the same as in the complete sample, we assessed whether the effect size of the association of the PDIs with physical and mental component scores changed after adding hsCRP to the model.

We observed that the effect sizes of PDI for the association with physical and mental well-being remained virtually unchanged (Table 5 and 6, Fig. 1). Accordingly, the association between the PDIs and well-being was not mediated by hsCRP, rather we observe additive effects of hsCRP and PDIs, suggesting that hsCRP levels are independently associated with physical but not mental well-being in older adults.

**Table 5.** Multivariate associations of a healthful plant-based diet (hPDI) and hsCRP with physical (PCS) and mental (MCS) well-being with hsCRP as second covariate of interest.

|  | OR [95%CI] | p |
| --- | --- | --- |
| **PCS** |  |  |
| **older adults (n=2251)** |  |  |
| hPDI medium | 1.25 [1.01; 1.56] | 0.039 |
| hPDI high | 1.12 [0.91; 1.38] | 0.287 |
| CRP | 0.97 [0.95; 1.00] | 0.026 |
| **younger adults (n=14699)** |  |  |
| hPDI medium | 1.09 [1.00; 1.18] | 0.041 |
| hPDI high | 1.15 [1.05; 1.25] | 0.003 |
| CRP | 0.98 [0.98; 1.00] | 0.017 |
| **MCS** |  |  |
| **older** |  |  |
| hPDI medium | 1.00 [0.81; 1.23] | 0.999 |
| hPDI high | 1.06 [0.87; 1.31] | 0.548 |
| CRP | 1.00 [0.98; 1.02] | 0.996 |
| **younger** |  |  |
| hPDI medium | 1.05 [0.97; 1.13] | 0.252 |
| hPDI high | 1.08 [0.99; 1.18] | 0.068 |
| CRP | 1.00 [1.00; 1.01] | 0.313 |

Adjusted for age, gender, BMI, physical activity, medication, income and energy intake

**Table 6.**
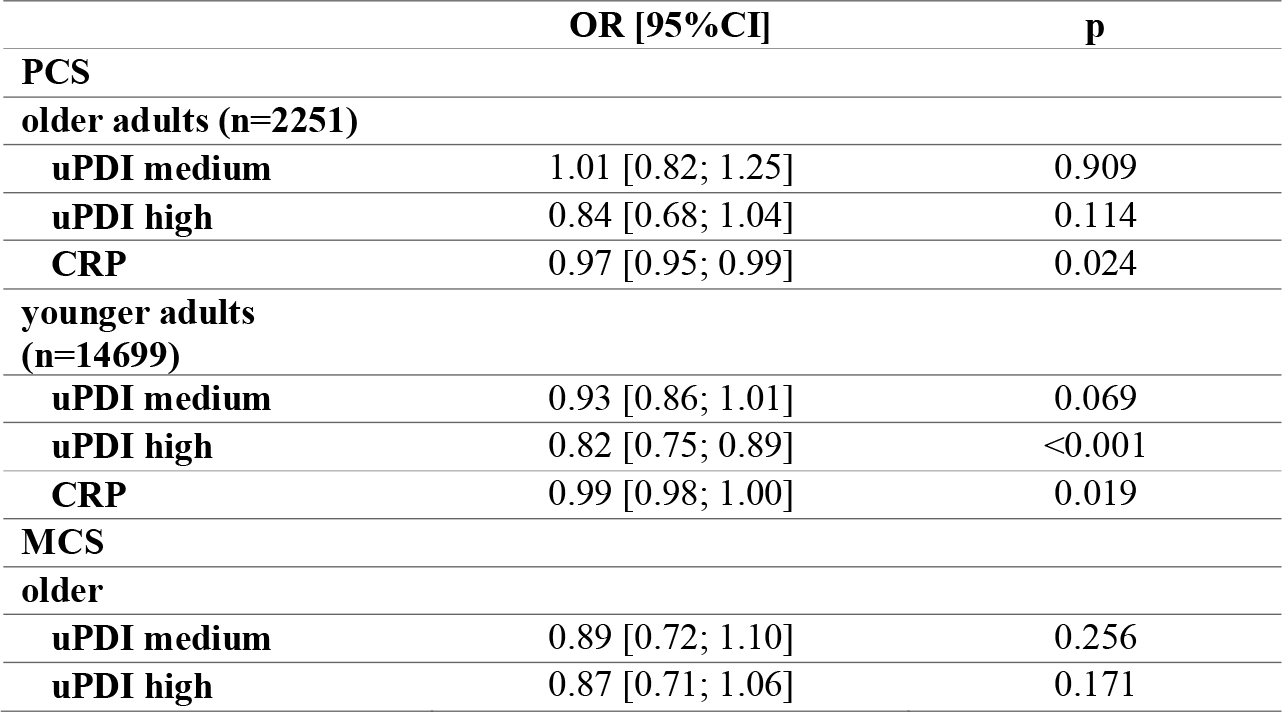
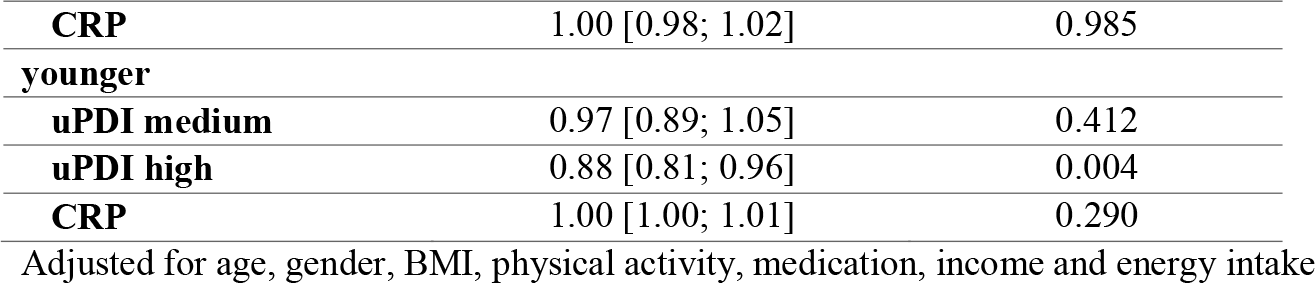
Multivariate associations of a unhealthful plant-based diet (uPDI) and hsCRP with physical (PCS) and mental (MCS) well-being with hsCRP as second covariate of interest.

**Fig. 1:**
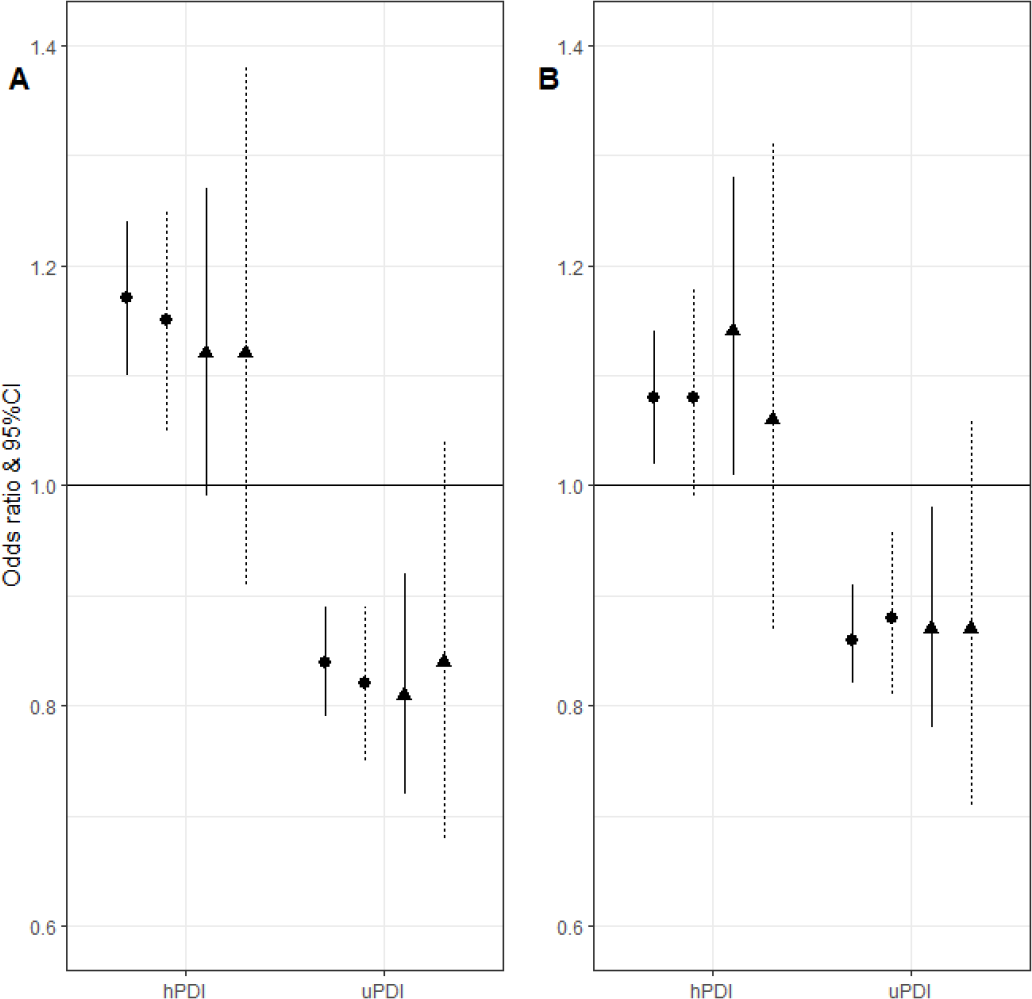
Associations between a healthful (hPDI) and unhealthful (uPDI) plant-based diet and well-being. A: association with physical well-being, B: association with mental well-being, •adults <60, adults >60 years, –model without hsCRP, ---model with hsCRP (Figure created in R-4.04.)

In younger adults, data on hsCRP were available in n=14699. In this subgroup, the associations between healthful and unhealthful plant-based diet index and well-being scores remained significant. For hPDI effect sizes changed in the smaller data set, but were not further affected by the addition of hsCRP. The addition of hsCRP to the regression model did not seem to influence the association between uPDI and physical and mental well-being, indicating independent effects (Table 6, Fig. 1). In both younger and older adults, hsCRP was negatively associated with physical but not with mental well-being (Table 5 and 6).

### Additional Analysis

As the majority of the population is more likely to follow an overall plant-based diet including a mixture of healthful and unhealthful plant-based food, we carried out a sensitivity analysis with the overall plant-based diet index, to assess how intake of plant foods in general associates with well-being. We observed a significant association between the highest tertile of PDI and physical well-being only among younger adults (OR 1.07, p=0.016), but no other significant association was observed (Suppl. Table 6).

Further, we replicated the analysis with a linear regression model with rank-inverse normal transformed component scores as the outcome. Our results show overall similar results, with a slight decrease in effect size in the smaller sample, however no mediating effect of hsCRP (Suppl. Table 7). Similarly, hsCRP was associated with physical and not mental well-being in these models.

## Discussion

In this study, we find high physical and mental well-being to be associated with high adherence to healthful plant-based diet. Similarly, low physical and mental well-being is associated with high adherence to unhealthful plan-based diet. We note slight differences between genders, but overlapping confidence intervals point towards no significant differences between males and females. Further, although age was a significant covariate in the models (Suppl. Table 8), our findings are largely similar for adults below and above 60 years. The main difference lies in the narrower confidence intervals among the younger age group, presumably due to greater sample size. Overall, our findings suggest that the association between plant-based diet adherence and physical and mental well-being does not significantly differ by age or sex. Inflammation, as measured by circulating hsCRP levels had no effect on the association between diet adherence and physical and mental well-being, indicating independent effects.

Our findings on the association between plant-based diet adherence and well-being, although cross-sectional, are consistent with earlier results [7]. Furthermore, our observation that hsCRP was significantly associated with physical, but not mental well-being, is mainly in line with previous observations [30, 31]. We extended previous research by assessing the association in men and women above 60 years of age. Further, we investigate state of inflammation as a potential mechanism explaining the association between plant-based diet adherence and well-being. The interplay of diet, inflammation and well-being was not assessed previously. By demonstrating that chronic inflammation levels and adherence to healthful plant-based diets are independently associated with physical and mental well-being, we have elucidated some of the factors contributing to well-being of older people and thereby likely to better survival into old age [17].

Since we found no mediating effect of hsCRP, an alternative explanation for the association between plant-based diets and well-being may lie in the gut microbiome. Previously, a link between gut microbial features and quality of life and depression has been observed [32, 33]. Although causality could so far not be established – i.e. whether depression led to changes in the gut microbiome or vice versa – the impact of diet on microbial composition has been shown in the past. Metabolites from protein degradation in the gut could have an indirect or direct effect on brain homeostasis, e.g. via production of neurotransmitters and vagal nerve stimulation, and consequently on mood [33, 34].

Short chain fatty acids, a product of microbial fiber fermentation in the gut, could directly have an impact on brain homeostasis or indirectly via activation of vagal nerves and stimulation of hormone production [33, 35]. Further research into the effects of plant-based diets on the gut microbiome is needed for understanding how this relation may impact well-being in older adults.

Particularly a healthful plant-based diet has been associated with increased diversity and anti-inflammatory features of the gut microbiome [36, 37]. Inflammation, a factor commonly assumed to play a role in the gut-brain axis, was however not a significant mediator in our analysis. An explanation may lie in our choice of inflammatory marker, as hsCRP is only one of many markers that can indicate inflammation. Previous research suggested that, for example glycoprotein acetyls may be a more sensitive marker for metabolic disease and gut microbiome function [38]. Apart from that, IL-6, as a marker of inflammation, has been found to be weakly but negatively associated with well-being [16]. These markers were not available to us, but future studies may explore an interaction with novel markers of inflammation.

This study also has some important limitations. The study population was overall relatively healthy and had high well-being scores compared to Dutch norms [26]. Also levels of circulating hsCRP were only slightly elevated among the older age group compared to the younger age group (Table 1, Suppl. Table 2), indicating an overall good health among older adults in the sample. This may have led to low variation and difficulty detecting effects of elevated hsCRP levels. The mean age of our older age group of 65 years is also still relatively young. As critical aspects of plant-based diets, such as energy deficits, may only become a problem among very old adults, and also inflammation increases with age, repeating the analysis in this age group may have led to different results. Secondly, we did not have data on vitamin D levels of participants available, although previous studies suggested an association of vitamin D levels with well-being [39, 40]. Further, we chose a logistic regression for our analysis, for which well-being scores had to be binarized. The arbitrary cut-off at the median may be criticized, however a sensitivity analysis conducted by linear regression with normal-transformed component scores led to overall similar results (Suppl. Table 7). Lastly, this analysis was cross-sectional, meaning causality cannot be inferred. Accordingly, reverse causation could contribute to the findings. Previous research showed that stress and depressive symptoms can lead to less healthy food choices and have associated depressive symptoms to poorer diet quality [41, 42]. However, meta-analysis of longitudinal studies suggests, that a high quality diet, may have a somewhat protective effect on depression risk [43].

Strengths of our analysis include the large sample size from the population-representative Lifelines Cohort Study. Secondly, hsCRP is a widely used measure for inflammation, allowing for comparison with previously publications. The RAND-36 is a commonly used tool to assess well-being and has also been validated in a Dutch population [44]. The size and abundance of the Lifelines study allowed us to assess the interplay between plant-based diets, well-being and inflammation in an age- and sex-diverse sample.

## Conclusion

In conclusion, we found a healthful plant-based diet to be associated with greater physical and mental well-being in an age- and sex-diverse population, whereas the opposite association was found for adherence to an unhealthful plant-based diet. This effect was not mediated by circulating levels of hsCRP, while this inflammatory marker was independently associated with lower physical well-being. A healthful plant-based diet may therefore contribute to sustain well-being in older ages. Future studies may focus on mechanisms that explain how plant-based diets may impact well-being via the gut microbiome or novel inflammatory markers.

## Data Availability

Data may be obtained from a third party and are not publicly available. Researchers can apply to use the Lifelines data used in this study. More information about how to request Lifelines data and the conditions of use can be found on their website (https://www.lifelines.nl/researcher/how-to-apply).

## Acknowledgements

The authors wish to acknowledge the services of the Lifelines Cohort Study, the contributing research centres delivering data to Lifelines, and all the study participants. The Lifelines initiative has been made possible by subsidy from the Dutch Ministry of Health, Welfare and Sport, the Dutch Ministry of Economic Affairs, the University Medical Center Groningen (UMCG), Groningen University and the Provinces in the North of the Netherlands (Drenthe, Friesland, Groningen).

This work has received funding from the European Union’s Horizon 2020 research and innovation framework under the Marie Sklodowska-Curie grant agreement No 860173. P.E. Slagboom, M. Beekman and L.C.P.G.M de Groot have received funding from the Vitality Oriented Innovations for the Lifecourse of the Ageing Society (VOILA) Consortium (ZonMw 457001001).

## Statements and Declarations

### Author Contribution

All authors contributed to the study conception and design. Material preparation and analysis were performed by Kerstin Schorr. The first draft of the manuscript was written by Kerstin Schorr and all authors commented on previous versions of the manuscript. All authors read and approved the final manuscript.

### Ethics approval

This study was performed in line with the principles of the Declaration of Helsinki. Approval was granted by the UMCG Ethics Committee (2007/152).

### Consent to participate

Informed consent was obtained from all individual participants included in the study.

## Footnotes

1 https://www.voedingscentrum.nl/nl/gezond-eten-met-de-schijf-van-vijf/wat-staat-in-de-schijf-van-vijf-en-wat-niet/olien-en-vetten.aspx

2 https://www.rand.org/health-care/surveys_tools/mos/36-item-short-form/scoring.html

