## Supplemental Material for "A healthful plant-based diet is associated with higher physical and mental well-being among older adults independent of circulating CRP"

MB: 0000-0003-0585-6206

PES: 0000-0002-2875-4723

CPGMdG: 0000-0003-2778-2789

JHMV: 0000-0001-6495-4207

Keywords: plant-based diet, subjective well-being, older adults, inflammation, dietary pattern

**Acknowledgements**

The authors wish to acknowledge the services of the Lifelines Cohort Study, the contributing research centres delivering data to Lifelines, and all the study participants. The Lifelines initiative has been made possible by subsidy from the Dutch Ministry of Health, Welfare and Sport, the Dutch Ministry of Economic Affairs, the University Medical Center Groningen (UMCG), Groningen University and the Provinces in the North of the Netherlands (Drenthe, Friesland, Groningen)

**Funding**

This work has received funding from the European Union’s Horizon 2020 research and innovation framework under the Marie Sklodowska-Curie grant agreement No 860173. P.E. Slagboom, M. Beekman and L.C.P.G.M de Groot have received funding from the Vitality Oriented Innovations for the Lifecourse of the Ageing Society (VOILA) Consortium (ZonMw 457001001).

**Supplementary Material**

**Suppl. Table 1 Categorization of food items and scoring of the PDI [19]**

| *Plant food groups* |  |
| --- | --- |
| Healthy |  |
| Vegetables | Fresh vegetables, lettuce, potatoes without fat |
| Fruits | fresh fruit |
| Nuts | Nuts, peanut butter |
| Legumes | Legumes, soup with legumes, soy-based meat alternatives, soymilk |
| Wholegrains | Brown and wholegrain bread and breadrolls, wholegrain pasta and rice |
| Vegetable Oils | Vegetable oils used for cooking, oil-based salad dressings, margarines |
| Tea & Coffee | Tea and coffee including decaffeinated coffee |
| *Less healthy* |  |
| Refined grains | White bread, rice and pasta, crackers, chips, cruesli, raisin bread |
| Fruit Juices | Fruit juices and fruit drinks |
| Potatoes | Fried potatoes, fries, kroketten, potatoes prepared with butter/margarine |
| Sweets & Desserts | Chocolate sprinkles, syrup, sugar added to coffee, cakes/biscuits, tarts, candybars, chocolate |
| Sugar sweetened beverages | Soda |
| *Animal food groups* |  |
| Animal fats | Butter, lard |
| Eggs | Eggs |
| Dairy | Milk, including milk in coffee, cheese, ice cream, yogurt |
| Fish | Dark meat fish, other fish, seafood, fried fish, salads with fish |
| Meat & Meatproducts | Chicken, pork, beef, bacon, sausages, burgers, offal |
| Miscellaneous animal-based foods | Pizza, salades, kroket |

**Suppl. Material 1: Overview over regression models**

Model 1 (in 4 groups stratified by gender and age above/below 60 years):

a) Mental Component Score as outcome, hPDI as predictor
b) Mental Component Score as outcome, uPDI as predictor
c) Physical Component Score as outcome, hPDI as predictor
d) Physical Component Score as outcome uPDI as predictor

Model 2 (in 4 groups stratified by gender and age above/below 60 years):XXXX

a) Mental Component Score as outcome, hPDI as predictor
b) Mental Component Score as outcome, uPDI as predictor
c) Physical Component Score as outcome, hPDI as predictor
d) Physical Component Score as outcome uPDI as predictor

Model 3 (in 4 groups stratified by gender and age above/below 60 years):XXXX

a) Mental Component Score as outcome, overall PDI as predictor
b) Physical Component Score as outcome overall PDI as predictor

**Suppl. Table 2: Characteristics of younger population (<60 years)**

|  | uPDI | | | hpdi | | |
| --- | --- | --- | --- | --- | --- | --- |
|  | **low** | **medium** | **high** | **low** | **medium** | **high** |
| **hPDI [18-90]** | 57.8±6.4 | 53.3±6.2 | 49.0±6.1 | 45.9±3.5 | 54.0±2.0 | 62.2±3.8 |
| **uPDI [18-90]** | 46.9±3.5 | 55.0±2.0 | 63.0±3.6 | 58.8±6.3 | 54.2±6.2 | 50.1±6.1 |
| **n** | 12199 | 13212 | 10439 | 12261 | 13205 | 10384 |
| **n_females_ (%)** | 7471 (61.2) | 7692 (58.2) | 6446 (61.7) | 7409 (60.4) | 7961 (60.3) | 6239 (60.1) |
| **Age, mean±SD** | 45.9 ± 8.8 | 42.6±9.5 | 38.5±10.0 | 39.8±10.1 | 43.0±9.6 | 45.1±9.1 |
| **BMI [kg/m^2^], mean±SD** | 25.7±3.8 | 25.4±3.9 | 25.0±4.1 | 25.5±4.1 | 25.5±4.0 | 25.2±4.0 |
| **Physical Activity (h/week), mean±SD** | 8.2±10.2 | 8.1±10.6 | 8.7±11.6 | 8.0±10.7 | 8.2±10.6 | 8.8±10.9 |
| **Medication use yes, n (%)** | 5361 (43.9) | 5640 (42.7) | 4556 (43.6) | 5392 (44.0) | 5694 (43.1) | 4471 (43.1) |
| **Monthly income, n (%)** |  |  |  |  |  |  |
| **(<1500€)** | 1266 (10.4) | 1742 (13.2) | 2110 (20.2) | 2138 (17.4) | 1704 (12.9) | 1276 (12.3) |
| **(1500-3000€)** | 5517 (45.2) | 6360 (48.1) | 5142 (49.3) | 5719 (46.6) | 6323 (47.9) | 4977 (47.9) |
| **(>3000€)** | 5416 (44.4) | 5110 (38.7) | 3187 (30.5) | 4431 (36.1) | 5162 (39.1) | 4120 (39.7) |
| **PCS, mean±SD** | 53.6±7.0 | 53.7±6.9 | 53.7±6.8 | 53.6±7.0 | 53.7±6.9 | 53.8±6.8 |
| **MCS, mean±SD** | 51.3±8.0 | 50.8±8.2 | 50.2±8.5 | 50.5±8.3 | 50.9±8.1 | 51.0±8.2 |
| **Including hsCRP (n=14699)** | | | | | | |
| **hsCRP [mg/L],**  **mean±SD** | 2.2±3.6 | 2.2±3.9 | 2.5±4.4 | 2.6±4.4 | 2.2±3.9 | 2.1±3.5 |

hPDI=healthy plant-based diet index, uPDI=unhealthful plant-based diet index, BMI=body mass index, PCS=physical component score, MCS=mental component score, hsCRP=high-sensitivity c-reactive protein

**Suppl. Table 3: Nutrient intake by tertile of hPDI and uPDI (>60 years)**

|  | uPDI | | | hPDI | | |
| --- | --- | --- | --- | --- | --- | --- |
|  | low | medium | high | low | medium | high |
| Energy [kcal/d] | 2015.0± 482.5 | 2005.9±483.2 | 2053.2±524.4 | 1941.7±474.4 | 2002.8±482.1 | 2129.2±513.4 |
| Carbo-hydrates [g/d] | 206.7±53.4 | 212.4±54.2 | 226.2±61.5 | 204.7±54.1 | 213.0±55.6 | 225.7±58.7 |
| Sugar [g/d] | 90.3±28.3 | 98.1±31.6 | 109.5±39.0 | 98.5±34.5 | 99.2±34.1 | 97.9±32.7 |
| Fiber [g/d] | 24.7±6.2 | 22.8±6.0 | 21.4±6.2 | 19.8±5.0 | 22.9±5.4 | 26.7±6.5 |
| Fats [g/d] | 82.1±28.2 | 82.5±28.8 | 84.2±30.6 | 80.1±27.4 | 81.2±28.4 | 87.2±31.0 |
| MUFA [g/d] | 28.5±10.5 | 28.4±10.9 | 28.5±11.0 | 27.5±10.0 | 28.0±10.4 | 30.1±11.8 |
| PUFA [g/d] | 17.9±8.2 | 17.5±8.3 | 17.6±9.0 | 15.7±7.0 | 17.3±8.0 | 20.2±9.6 |
| SFA [g/d] | 28.6±10.0 | 29.5±10.3 | 30.9±11.3 | 30.0±10.8 | 29.0±10.5 | 29.7±10.4 |
| DHA [g/d] | 0.14±0.1 | 0.11±0.1 | 0.09±0.1 | 0.12±0.1 | 0.12±0.1 | 0.11±0.1 |
| EPA [g/d] | 0.1±0.1 | 0.08±0.1 | 0.06±0.1 | 0.8±0.1 | 0.8±0.1 | 0.8+0.1 |
| Protein [g/d] | 81.3±16.7 | 75.4±15.8 | 71.8±16.9 | 73.7±16.0 | 76.2±16.5 | 80.0±17.7 |
| Plant protein [g/d] | 32.8±9.2 | 30.7±8.7 | 29.6±9.0 | 27.0±7.1 | 30.7±7.9 | 36.0±9.7 |
| Animal protein [g/d] | 48.5±12.8 | 44.7±12.2 | 42.3±12.4 | 46.8±12.3 | 45.6±12.5 | 44.0±13.4 |
| Vitamin B12 [mcg/d] | 5.0±2.5 | 4.5±2.2 | 4.1±2.1 | 4.9±2.5 | 4.5±2.2 | 4.2±2.2 |

MUFA: monounsaturated fatty acids, PUFA: polyunsaturated fatty acids, SFA: saturated fatty acids, DHA: docosahexaenoic acid, EPA: eicosapentaenoic acid

**Suppl. Table 4 Association between an unhealthful plant-based diet index (uPDI) and physical (PCS) and mental (MCS) well-being, stratified by gender among younger adults**

|  | Physical component score | | Mental component score | |
| --- | --- | --- | --- | --- |
| Females | OR [95% CI] | p | OR [95% CI] | p |
| hPDI Tertile 1 |  |  |  |  |
| hPDI Tertile 2 | 1.10 [1.03;1.18] | 0.005 | 1.00 [0.94; 1.07] | 0.960 |
| hPDI Tertile 3 | 1.10 [1.02,1.19] | 0.009 | 1.03 [0.96; 1.11] | 0.400 |
| Males |  |  |  |  |
| hPDI Tertile 1 |  |  |  |  |
| hPDI Tertile 2 | 1.12 [1.03; 1.21] | 0.007 | 1.07 [0.99; 1.16] | 0.079 |
| hPDI Tertile 3 | 1.27 [1.16; 1.39] | <0.001 | 1.12 [1.03; 1.22] | 0.010 |
| Females |  |  |  |  |
| uPDI Tertile 1 | ref |  | ref |  |
| uPDI Tertile 2 | 0.95 [0.89; 1.01] | 0.113 | 0.92 [0.86; 0.99] | 0.016 |
| uPDI Tertile 3 | 0.87 [0.81; 0.93] | <0.001 | 0.89 [0.82; 0.95] | <0.001 |
| Males |  |  |  |  |
| uPDI Tertile 1 | ref |  | ref |  |
| uPDI Tertile 2 | 0.85 [0.78; 0.93] | <0.001 | 0.96 [0.89; 1.05] | 0.390 |
| uPDI Tertile 3 | 0.81 [0.75; 0.88] | <0.001 | 0.88 [0.81; 0.95] | 0.002 |

adjusted for age, BMI, physical activity, income and medication use

**Suppl. Table 5: Mediation analysis**

|  | Beta [95%CI] | p |
| --- | --- | --- |
| Older adults |  |  |
| uPDI medium | 0.03 [-0.07; 0.12] | 0.583 |
| uPDI high | 0.07 [-0.03; 0.16] | 0.172 |
| hPDI medium | -0.15 [-0.24; -0.05] | 0.003 |
| hPDI high | -0.09 [-0.18; 0.00] | 0.057 |
| Younger adults |  |  |
| uPDI medium | 0.03 [-0.01; 0.06] | 0.117 |
| uPDI high | 0.12 [0.08; 0.15] | <0.001 |
| hPDI medium | -0.08 [-0.12; -0.05] | <0.001 |
| hPDI high | -0.12 [-0.16; -0.09] | <0.001 |

**Suppl. Table 6 Association of an overall plant-based diet index (PDI) with physical (PCS) and mental (MCS) component score**

|  | Physical component score | | Mental component score | |
| --- | --- | --- | --- | --- |
| Older adults | OR [95% CI] | p | OR [95% CI] | p |
| Tertile 1 | ref |  | ref |  |
| Tertile 2 | 0.98 [0.87; 1.11] | 0.755 | 0.90 [0.80; 1.02] | 0.098 |
| Tertile 3 | 1.02 [0.90; 1.15] | 0.773 | 0.94 [0.84; 1.07] | 0.352 |
| Younger adults |  |  |  |  |
| Tertile 1 | ref |  | ref |  |
| Tertile 2 | 1.02 [0.97; 1.07] | 0.416 | 0.98 [0.93; 1.03] | 0.320 |
| Tertile 3 | 1.07 [1.01; 1.13] | 0.016 | 0.91 [0.91; 1.01] | 0.130 |

adjusted for age, gender, BMI, physical activity, income, medication use and energy intake

**Suppl. Table 7 Association between a healthful (hPDI) or unhealthful (uPDI) plant-based diet and physcial (PCS) and mental (MCS) well-being assessed by linear regression with rank-inverse normal transformation**

| Model 1 | PCS  Beta [95%CI] | p | MCS  Beta [95%CI] | p |
| --- | --- | --- | --- | --- |
| Older adults |  |  |  |  |
| uPDI | -0.006 [-0.01; -0.002] | 0.002 | -0.006 [-0.01; 0.002] | 0.002 |
| hPDI | 0.003 [0.00; 0.007] | 0.056 | 0.003 [0.00; 0.007] | 0.078 |
| Younger adults |  |  |  |  |
| uPDI | -0.006 [-0.007; -0.004] | <0.001 | -0.004 [-0.006; -0.003] | <0.001 |
| hPDI | 0.005 [0.004; 0.007] | <0.001 | 0.003 [0.001; 0.004] | <0.001 |
| Model 2 |  |  |  |  |
| Older adults |  |  |  |  |
| uPDI | -0.005 [-0.011, 0.002] | 0.159 | -0.002 [-0.009; 0.004] | 0.482 |
| hPDI | 0.004 [-0.002; 0.01] | 0.212 | 0.001 [-0.005; 0.008] | 0.687 |
| Younger adults |  |  |  |  |
| uPDI | -0.005 [-0.007;-0.003] | <0.001 | -0.004 [-0.007; -0.002] | <0.001 |
| hPDI | 0.005 [0.003; 0.007] | <0.001 | 0.003 [0.00; 0.005] | 0.022 |
| Model 3 |  |  |  |  |
| Older adults |  |  |  |  |
| uPDI | -0.004 [-0.011; 0.002] | 0.168 | -0.002 [-0.009; 0.004] | 0.480 |
| hPDI | 0.004 [-0.002; 0.01] | 0.230 | 0.001 [-0.005; 0.008] | 0.683 |
| Younger adults |  |  |  |  |
| uPDI | -0.005 [-0.007; -0.003] | <0.001 | -0.004 [-0.007; -0.002] | <0.001 |
| hPDI | 0.005 [0.003; 0.007] | <0.001 | 0.003 [0.000; 0.005] | 0.022 |

Model 1: adjusted for age, gender, bmi, physical activity, income, medication use and energy intake in subsample (n_>60_=6635, n_<60_=35850);

Model 2: adjusted for age, gender, bmi, physical activity, income, medication use and energy intake in subsample (n_>60_=2251, n_<60_=14699);

Model 3: adjusted for age, gender, bmi, physical activity, income, medication use energy intake and hsCRP in subsample (n_>60_=2251, n_<60_=14699)

**Suppl. Table 8: Effect sizes of covariates in the different models**

|  | hPDI | | | | uPDI | | | |
| --- | --- | --- | --- | --- | --- | --- | --- | --- |
|  | **PCS** | | **MCS** | | **PCS** | | **MCS** | |
|  | **OR [95% CI]** | **p** | **OR [95% CI]** | **p** | **OR [95% CI]** | **p** | **OR [95% CI]** | **p** |
| Older Adults |  |  |  |  |  |  |  |  |
| Model 1 |  |  |  |  |  |  |  |  |
| age | 0.98 [0.97; 0.99] | <0.001 | 1.02 [1.01; 1.03] | <0.001 | 0.98 [0.97;0.99] | <0.001 | 1.02 [1.01; 1.03] | <0.001 |
| Gender | 1.53 [1.37; 1.72] | <0.001 | 1.87 [1.67; 2.09] | <0.001 | 1.53 [1.36; 1.72] | <0.001 | 1.85 [1.65; 2.07] | <0.001 |
| Physical Activity | 1.02 [1.01; 1.02] | <0.001 | 1.01 [1.00; 1.01] | 0.07 | 1.02 [1.01; 1.02] | <0.001 | 1.01 [1.00; 1.01] | 0.071 |
| BMI | 0.92 [0.90; 0.93] | <0.001 | 1.03 [1.02; 1.05] | <0.001 | 0.92 [0.90;0.93] | <0.001 | 1.03 [1.02; 1.05] | <0.001 |
| Income | 1.19 [1.10; 1.29] | <0.001 | 1.24 [1.14; 1.34] | <0.001 | 1.18 [1.09; 1.28] | <0.001 | 1.23 [1.14; 1.33] | <0.001 |
| Medication | 2.12 [1.91; 2.37] | <0.001 | 1.19 [1.07; 1.32] | 0.001 | 2.13 [1.91; 2.37] | <0.001 | 1.19 [1.07; 1.32] | 0.001 |
| Energy intake | 1.00 [1.00; 1.00] | <0.001 | 1.00 [1.00; 1.00] | <0.001 | 1.00 [1.00; 1.00] | 0.001 | 1.00 [1.00; 1.00] | <0.001 |
| Model 2 |  |  |  |  |  |  |  |  |
| age | 0.97 [0.95; 0.99] | 0.002 | 1.02 [1.01; 1.04] | 0.011 | 0.97 [0.95; 0.99] | 0.002 | 1.02 [1.01; 1.04] | 0.010 |
| Gender | 1.55 [1.28; 1.89] | <0.001 | 1.99 [1.64; 2.40] | <0.001 | 1.53 [1.26; 1.86] | <0.001 | 2.00 [1.65; 2.41] | <0.001 |
| Physical Activity | 1.02 [1.00; 1.03] | 0.006 | 1.00 [0.99; 1.01] | 0.694 | 1.02 [1.00; 1.03] | 0.007 | 1.00 [0.99; 1.01] | 0.663 |
| BMI | 0.93 [0.91; 0.96] | <0.001 | 1.02 [1.00; 1.05] | 0.048 | 0.93 [0.91; 0.96] | <0.001 | 1.02 [1.00; 1.05] | 0.063 |
| Income | 1.27 [1.10; 1.45] | <0.001 | 1.19 [1.04; 1.36] | 0.010 | 1.25 [1.09; 1.44] | 0.001 | 1.18 [1.03; 1.35] | 0.016 |
| Medication | 2.05 [1.70; 2.46] | <0.001 | 1.06 [0.89; 1.27] | 0.50 | 2.05 [1.71; 2.47] | <0.001 | 1.06 [0.89; 1.27] | 0.50 |
| Energy intake | 1.00 [1.00; 1.00] | 0.101 | 1.00 [1.00; 1.00] | 0.007 | 1.00 [1.00; 1.00] | 0.153 | 1.00 [1.00; 1.00] | 0.008 |
| hsCRP | 0.97 [0.95; 1.00] | 0.026 | 1.00 [0.98; 1.02] | 0.996 | 0.97 [0.95; 1.00] | 0.024 | 1.00 [0.98; 1.02 | 0.985 |
| Younger Adults |  |  |  |  |  |  |  |  |
| Model 1 |  |  |  |  |  |  |  |  |
| age | 0.97 [0.97; 0.98] | <0.001 | 1.03 [1.01; 1.01] | <0.001 | 0.97 [0.97; 0.98] | <0.001 | 1.01 [1.01; 1.01] | <0.001 |
| Gender | 1.23 [1.17; 1.29] | <0.001 | 1.62 [1.54; 1.70] | <0.001 | 1.21 [1.15; 1.28] | <0.001 | 1.61 [1.53; 1.69] | <0.001 |
| Physical Activity | 1.00 [1.00; 1.01] | 0.005 | 1.01 [1.00; 1.01] | <0.001 | 1.00 [1.00; 1.01] | 0.004 | 1.01 [1.00; 1.01] | <0.001 |
| BMI | 0.93 [0.92; 0.93] | <0.001 | 1.01 [1.01; 1.02] | <0.001 | 0.93 [0.92; 0.93] | <0.001 | 1.01 [1.01; 1.02] | <0.001 |
| Income | 1.17 [1.13; 1.20] | <0.001 | 1.28 [1.24; 1.32] | <0.001 | 1.16 [1.12; 1.20] | <0.001 | 1.27 [1.23; 1.31] | <0.001 |
| Medication | 1.82 [1.74; 1.90] | <0.001 | 1.30 [1.25; 1.36] | <0.001 | 1.82 [1.74; 1.90] | <0.001 | 1.30 [1.25; 1.36] | <0.001 |
| Energy intake | 1.00 [1.00; 1.00] | <0.001 | 1.00 [1.00; 1.00] | <0.001 | 1.00 [1.00; 1.00] | <0.001 | 1.00 [1.00; 1.00] | <0.001 |
| Model 2 |  |  |  |  |  |  |  |  |
| age | 0.97 [0.97; 0.98] | <0.001 | 1.01 [1.01; 1.01] | <0.001 | 0.97 [0.97; 0.98] | <0.001 | 1.01 [1.01; 1.01] | <0.001 |
| Gender | 1.15 [1.06; 1.24] | 0.001 | 1.61 [1.49; 1.75] | <0.001 | 1.13 [1.04; 1.23] | 0.003 | 1.60 [1.47; 1.73] | <0.001 |
| Physical Activity | 1.00 [1.00; 1.00] | 0.531 | 1.01 [1.00; 1.01] | <0.001 | 1.00 [1.00; 1.00] | 0.475 | 1.01 [1.00; 1.01] | <0.001 |
| BMI | 0.93 [0.92; 0.94] | <0.001 | 1.01 [1.00; 1.02] | 0.067 | 0.93 [0.92; 0.94] | <0.001 | 1.01 [1.00; 1.02] | 0.104 |
| Income | 1.15 [1.10; 1.21] | <0.001 | 1.28 [1.22; 1.34] | <0.001 | 1.14 [1.09; 1.20] | <0.001 | 1.27 [1.21; 1.34] | <0.001 |
| Medication | 1.82 [1.70; 1.95] | <0.001 | 1.30 [1.22; 1.39] | <0.001 | 1.82 [1.70; 1.95] | <0.001 | 1.30 [1.21; 1.39] | <0.001 |
| Energy intake | 1.00 [1.00; 1.00] | 0.135 | 1.00 [1.00; 1.00] | <0.001 | 1.00 [1.00; 1.00] | 0.419 | 1.00 [1.00; 1.00] | <0.001 |
| hsCRP | 0.99 [0.98; 1.00] | 0.017 | 1.00 [1.00; 1.01 | 0.313 | 0.99 [0.98; 1.00] | 0.019 | 1.00 [1.00; 1.01] | 0.290 |
